# Composite Deep Network with Feature Weighting for Improved Delineation of COVID Infection in Lung CT

**DOI:** 10.1101/2023.01.17.23284673

**Authors:** Pallabi Dutta, Sushmita Mitra

## Abstract

An early effective screening and grading of COVID-19 has become imperative towards optimizing the limited available resources of the medical facilities. An automated segmentation of the infected volumes in lung CT is expected to significantly aid in the diagnosis and care of patients. However, an accurate demarcation of lesions remains problematic due to their irregular structure and location(s) within the lung.

A novel deep learning architecture, Composite Deep network with Feature Weighting *(CDNetFW)*, is proposed for efficient delineation of infected regions from lung CT images. Initially a coarser-segmentation is performed directly at shallower levels, thereby facilitating discovery of robust and discriminatory characteristics in the hidden layers. The novel feature weighting module helps prioritise relevant feature maps to be probed, along with those regions containing crucial information within these maps. This is followed by estimating the severity of the disease.

The deep network *CDNetFW* has been shown to outperform several state-of-the-art architectures in the COVID-19 lesion segmentation task, as measured by experimental results on CT slices from publicly available datasets, especially when it comes to defining structures involving complex geometries.

## 1. Introduction

A critical step in the fight against COVID-19 is an effective screening of the level of infection in patients; such that those seriously affected can receive immediate treatment and care, as well as be isolated to mitigate the spread of the virus. The gold standard screening method currently used for detecting COVID-19 cases is the reverse transcription polymerase chain reaction (RT-PCR) testing, which is a very time-consuming, laborious, and complicated manual process. The test is uncomfortable, invasive, uses nasopharyngeal swabs, and has high false negative rates; with outcome being dependent on sampling errors and low viral load. Given that CT is reliable to check changes in the lungs, its importance in the context of COVID-19 becomes all the more evident.

Conspicuous ground-glass opacity (GGO) and multiple mottling lesions, in the peripheral and posterior lungs on CT images, are hallmark characteristics of COVID-19 pneumonia [1]. While the GGO are hazy darkened spots in the lung, diffuse enough such that they do not block underlying blood vessels or lung structures, the consolidations correspond to areas of increased lung density [1]. It is observed that with time these infection characteristics became more frequent, and are likely to spread across both lungs. Morozov *et al*. [2] suggested a partially quantitative severity grading system on a scale of 0 to 4, with a step value of 25%, based on the percentage of the diseased lung tissue. While CT-0 represents healthy cohorts, grade CT-4 refers to those patients with affected lung regions *>* 75%. This scale was assigned by experts based on visual inspection of the lung CT scans of infected patients.

In order to speed up the discovery of disease mechanisms, machine learning and deep learning [3] can be effectively employed to detect abnormalities and extract textural features of the altered lung parenchyma; to be subsequently related to specific signatures of the COVID-19 virus. Automated segmentation of the lung region and lesion from CT can help outline the volume of interest (VOI) for a fast detection and grading of the severity of infection. A consistent and reproducible method, for the rapid evaluation of high volumes of screening or diagnostic thoracic CT studies, is possible with artificial intelligence. Demarcation of affected lung tissues in the CT slices demands high precision. Deep learning enables circumventing the visual approximation by radiologists, to produce accurate decisions; particularly in the scenario of high volumes of disease cases.

Typically deep learning requires a machine to learn automatically from raw data to discover representations needed for detection or classification. In the context of medical images, it directly uses pixel values of the images at the input; thereby, overcoming the manual errors caused by inaccurate segmentation and/or subsequent hand-crafted feature extraction. Convolution neural networks (CNNs) [3] constitute one of the popular models of deep learning. Some of the commonly used deep learning models in medical applications include CNN, ResNet [4], Res2Net [5], DenseNet [6], and SegNet [7].

Custom architectures, like the *U*-Net [8], have been designed for segmentation with promising results. Here the skip connections between the symmetric down-sampling (encoder) and up-sampling (decoder) paths, in the fully convolutional framework, provide local (high resolution) information in the global context during up-sampling; thereby, resulting in improved localization of the target region with more precise segmentation results. However, it is observed that the target lesion regions (like GGO) in lung CT scans of COVID-19 patients are of lower contrast with blurred edges, particularly in the early stages. They also appear in varying shapes and sizes within the lung. Attention modules were incorporated in the vanilla *U*-Net framework [9], in order to enhance its segmentation performance. The encoder feature maps were recalibrated spatially with the aid of decoder feature maps. The attention module thus helped the network to focus only on the relevant regions of interest in the target, while suppressing the contribution from irrelevant areas.

Both ResNet and Res2Net incorporate residual connections, to help combat the vanishing gradient issue, while developing deeper networks. Res2Net additionally employs parallel cascading convolution operations within a block. This helps capture multi-scale features in the input volume. The DenseNet makes use of residual connections by bridging all convolution layers with each other. It additionally promotes re-use of feature maps, with further improvement in performance. The *U*-Net++ [10] and Residual *U*-Net [11] are also used in medical image segmentation. The *U*-Net++ inserts multiple convolution blocks between the encoding and decoding pathway of its *U−*Net backbone. This helps narrow down the semantic gap between the activation maps produced by the encoders and decoders; thereby, reducing the complexity of optimization. An exhaustive survey on the role of artificial intelligence in analysing and predicting COVID-19 can be found in [12, 13].

A novel deep network Composite Deep network with Feature Weighting (*CDNetFW*) is developed for efficiently demarcating the COVID-19 infection in lung CT images. It is capable of learning generalized representations of the irregularly structured infected regions. The contribution of this research is summarized below.

- A mini-segmentation architecture is introduced in the encoder branch of the composite deep network. Such coarser-level segmentation provides direct supervision to intermediate encoder layers for circumventing noisy activation and vanishing gradient.
- The feature weighting module focuses on the relevant activation responses. The activation maps carrying most pertinent data about the target regions are initially detected. This is followed by identification of the spatial locations-of-interest within them. Incorporation of depth-wise and dilated convolutions helps generate the relevant re-calibrating weights for map refinement. Dilated convolutions help capture multi-scale features towards improved segmentation of lesions having different sizes and textures.
- A combination loss is used to handle the issue of class imbalance, while simultaneously minimizing the occurrence of False Positives and False Negatives in the output.

The rest of the paper is organized as follows. Section 2 briefly describes state-of-the-art literature on the use of deep learning for segmentation of COVID lesions from lung CT images. In Section 3 we introduce the proposed *CDNetFW*. Experimental results are discussed in Section 4. Comparative study is provided with state-of-the-art models, including the *U−*Net [8] and its variants like *U−*Net++ [10], Residual *U*-Net [11], and Attention *U−*Net [9], on segmentation of the lesions. The demarcated region is used to compute the percentage of infection in the lung; on the basis of which a grading of the severity of the disease is evaluated. The superiority of the *CDNetFW* is demonstrated on CT slices extracted from four sets of publicly available data, *viz*. MOSMED [14], MedSeg-COV-1 [15], MedSeg-COV-2 [16], and COV-CT-Lung-Inf-Seg [17]. Finally Section 5 concludes the article.

## 2. Deep Learning for Delineating Infected CT Volumes

A retrospective, multi-centric study [18] employed the ResNet-50, with *U*-Net for segmenting the lung region from CT, to differentiate between COVID-19 viral pneumonia, Community acquired pneumonia (CAP) and other non-pneumonia images. Similarly, a 3D DenseNet was trained in a diverse multinational cohort [19] to localize parietal pleura/ lung parenchyma, followed by classification of COVID-19 pneumonia; with high sensitivity and specificity on an independent test set. Visualization of activation regions was performed using Grad-CAM to assess association of peripheral regions of the lung across variable amounts of disease burden. A novel Joint Classification and Segmentation system was designed [20] to perform real-time and explainable diagnosis of COVID-19 cases. Annotation was provided with fine-grained pixel-level labels, lesion counts, infected areas and locations, benefiting various diagnosis aspects. With the explainable classification outputs coming from Res2Net, and the corresponding fine-grained lesion segmentation resulting from VGG-16, the system helped simplify and accelerate the diagnostic process for radiologists.

An attention-based deep 3D multiple instance learning (AD3D-MIL) was developed [21] for the effective screening of COVID-19. A patient-level label was assigned to each 3D chest CT, which was viewed as a bag of instances. An attention-based pooling approach to 3D instances provided insight into the contribution of each instance towards the bag label.

The segmentation performance of the *U*-Net and SegNet was compared [22] on one out of the two datasets in MedSeg-29 [16], obtained from the Italian Society of Medical Interventional Radiology. While binary segmentation identified the lung tissues infected by COVID-19 virus, the multi-class segmentation delineated the different lesion pathologies. However, both exhibited poor performance in multi-class segmentation, particularly for identifying the pleural effusions, possibly due to scarcity of corresponding CT slices.

A multi-task architecture was introduced [23] to simultaneously perform classification and segmentation. The lung CT scans were grouped into COVID-19 infected and non-infected categories, while also ranking the infected scans in decreasing order of severity. The infected regions were first segmented to calculate the ratio of the affected lung region w.r.t. the total lung region, over each slice. The maximum value was used to compute the severity stage, as per the guidelines of the Fleischner Society.

The DUDA-Net [24] employed a cascaded pair of *U*-Nets, with the first one extracting the lung region from the CT slice to make it easier for subsequent focus on the COVID-19 lesions. Attention modules with dilated convolutions helped the second *U*-Net to effectively capture multi-scale information, and re-weight relevant channels of the input feature map volume for better focus on the region of interest. This strategy helped decrease any segmentation error caused by smaller-sized lesions. However this entailed an increase in computational complexity in terms of the parameters involved.

A pair of attention modules were introduced [25] in the *U*-Net framework, to enhance feature map volumes along the skip connections as well as the up-sampled feature maps of the decoder. This dual attention strategy helped re-weight the feature maps, both spatially and channel-wise, for improved segmentation of the infected lung tissues in the slices. The dilated convolutions helped generate larger receptive fields.

The nCoVSegNet [26] employed two-stage transfer learning to deal with data scarcity in large annotated COVID19 datasets. It acquired knowledge both from the ImageNet [27] and a lung nodule detection dataset (LIDC-IDRI), to delineate COVID-19 lesions from lung CT scans in the MOSMED dataset. A global context-aware module, employing convolution blocks of varying sizes, helped capture multi-scale features. A Dual-Attention fusion module, incorporating both channel and spatial attention, enabled improved segmentation performance.

The Inf-Net [28] involved a parallel partial decoder to aggregate high-level features for generating a global segmentation map. A reverse attention mechanism, with an additional edge attention module, was incorporated for better delineation of the blurred edges of the lesions. The semi-supervised framework, requiring only a few labeled images and leveraging primarily unlabeled data, helped improve the learning ability with a higher performance.

A lightweight CNN model LCOV-Net [29] employed a separable convolution operation, in lieu of the conventional convolution block, to capture features from 3D lung CT volumes; thereby, significantly reducing the model parameters to make it computationally lighter with faster training time. Attention mechanism was incorporated to recalibrate the feature maps, in order to emphasize the relevant features w.r.t. the region of interest.

## 3. Composite Deep Network with Feature Weighting (CDNetFW)

This section describes the *CDNetFW* along with its primary components, *viz*. the mini-segmentation network and the feature weighting mechanism. The loss function employed is also outlined, along with the performance evaluation metrics used.

### 3.1 Architecture

The architecture of *CDNetFW* is schematically depicted in Fig. 1. The CT slices are provided as input to a five-tier composite network. Unlike the conventional symmetric encoder-decoder framework used for segmentation, here the encoding path encompasses a mini-segmentation module with auxiliary decoder branches. Additional feature weighting modules are introduced to detect the relevant activation maps from the entire set, while identifying the spatial locations-of-interest within them. This helps reduce the computational burden. The detailed framework is summarized in Table 1.

**Figure 1:**
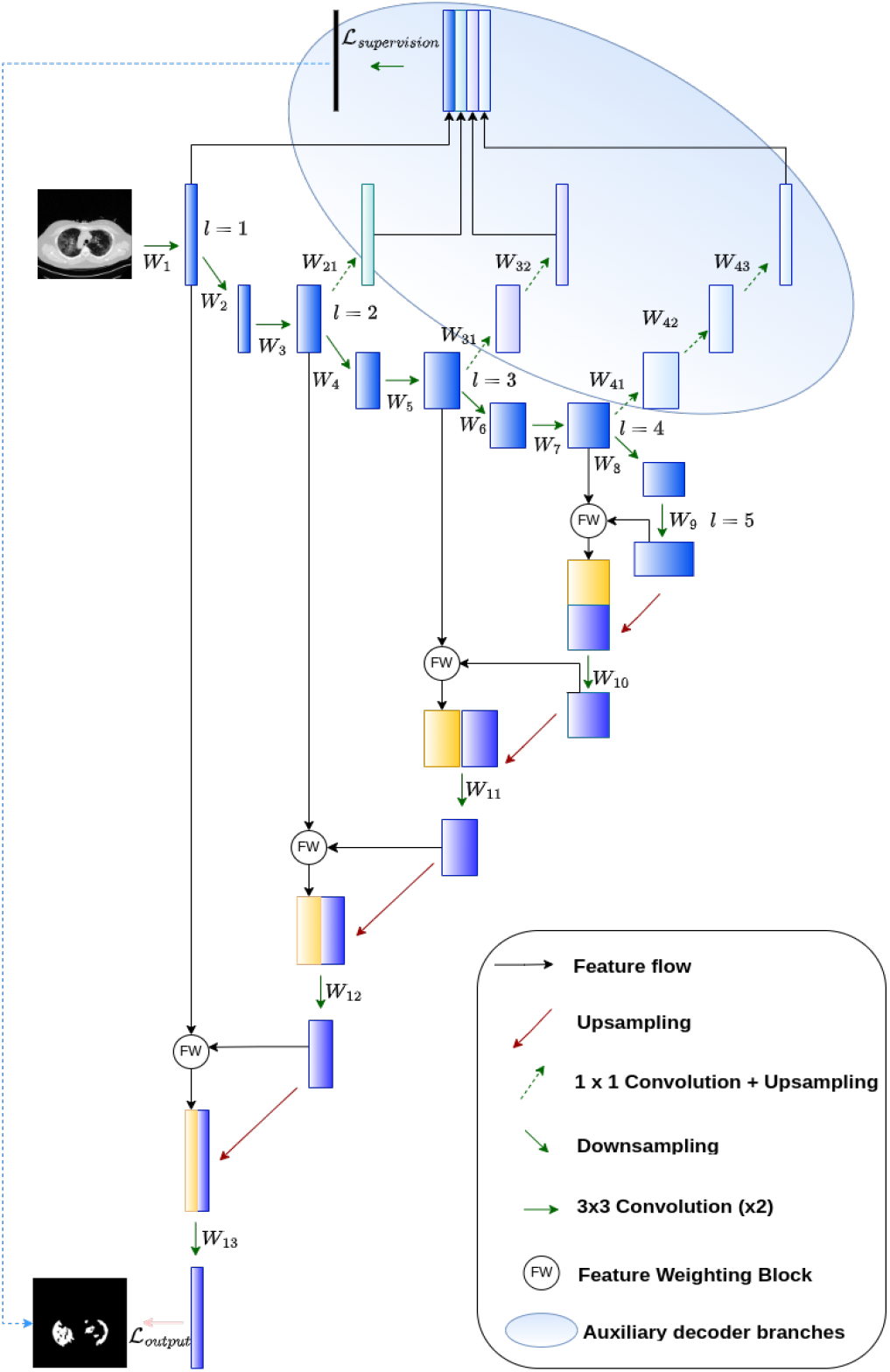
Schematic diagram of *CDNetFW*

**Table 1:** Architectural details of *CDNetFW*. Here Conv2D indicates 2D convolution, ZP represents zero padding, GN denotes Group Normalization, and UP2D corresponds to 2D upsampling with bilinear interpolation.

| Level | Encoder |  | Decoder |  | Deep Supervision |  |
| --- | --- | --- | --- | --- | --- | --- |
|  | Filters | Output | Filters | Output | Filters | Output |
| 1 | Input Layer | 512x512x1 | UP2D | 512x512x128 | N/A |  |
|  | Conv2D+ZP | 512x512x64 | Conv2D+ZP | 512x512x64 |  |  |
|  | GN | 512x512x64 | GN | 512x512x64 |  |  |
|  | Conv2D+ZP | 512x512x64 | Conv2D+ZP | 512x512x64 |  |  |
|  | Conv2D | 256x256x64 | Output Layer | 512x512x1 |  |  |
| 2 | Conv2D+ZP | 256x256x128 | UP2D | 256x256x256 | Conv2D+GN+UP2D | 512x512x64 |
|  | GN | 256x256x128 | Conv2D+ZP | 256x256x128 |  |  |
|  | Conv2D+ZP | 256x256x128 | GN | 256x256x128 |  |  |
|  | Conv2D | 128x128x128 | Conv2D+ZP | 256x256x128 |  |  |
| 3 | Conv2D+ZP | 128x128x256 | UP2D | 128x128x512 | (Conv2D+GN+UP2D) x2 | 512x512x64 |
|  | GN | 128x128x256 | Conv2D+ZP | 128x128x256 |  |  |
|  | Conv2D+ZP | 128x128x256 | GN | 128x128x256 |  |  |
|  | Conv2D | 64x64x256 | Conv2D+ZP | 128x128x256 |  |  |
| 4 | Conv2D+ZP | 64x64x512 | UP2D | 64x64x1024 | (Conv2D+GN+UP2D) x3 | 512x512x64 |
|  | GN | 64x64x512 | Conv2D+ZP | 64x64x512 |  |  |
|  | Conv2D+ZP | 64x64x512 | GN | 64x64x512 |  |  |
|  | Conv2D | 32x32x512 | Conv2D+ZP | 64x64x512 |  |  |
| 5 | Conv2D+ZP | 32x32x1024 | N/A |  | N/A |  |
|  | GN | 32x32x1024 |  |  |  |  |
|  | Conv2D+ZP | 32x32x1024 |  |  |  |  |

### 3.2 Mini-segmentation module

Image segmentation in deep networks typically employs a symmetric structure of encoding and decoding pathways [8, 9, 11]. While increasing the depth of the model enhances its prediction performance, the issues like noisy feature maps at shallower levels, vanishing gradients, overfitting, etc. remain. Although skip connections are included to tackle the problem of vanishing gradients, often less discriminative features remain prevalent (as observed from Fig. 7(c)). This leads to poorer generalisation capacity. In order to circumvent this problem, we propose to introduce supervision at shallower levels of the encoder through the embedding of a mini-segmentation network within it. Auxiliary decoder branches are attached to the second, third, and fourth levels of the encoder arm, as illustrated in Fig. 1. These auxiliary decoding paths provide additional supervision during training. Minimization of the loss function *ℒ*_*supervision*_ at the output of this mini-segmentation network aids in enhancing the quality of the output activation at each level of the encoder. The impact of vanishing gradient is reduced by superposing the gradients obtained from these auxiliary decoder branches with that at the final output layer.

Let Θ = [*θ*_1_, *θ*_2_] consist of *θ*_1_ = [*W*_21_, *W*_31_, *W*_32_, *W*_41_, *W*_42_, *W*_43_], representing the weights for the auxiliary deep supervision branches, and *θ*_2_ = [*W*_1_, …, *W*_7_], corresponding to the weights of the encoding path (Fig. 1). The adaptation is expressed as

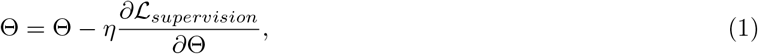

where *ℒ*_*supervision*_ is the auxiliary deep supervision loss and *η* is the learning rate.

### 3.3 Feature weighting module

Concatenating encoder activation maps with decoder feature representations, via skip connections, helps incorporate lower-level spatial information towards improved target localization. The *Feature Weighting (FW)* mechanism allows focus on the relevant activation regions. Simultaneous channel and spatial refinement, incorporated in *FW* block, enables better segmentation of the target lesions. While channel refinement highlights feature maps of interest in the input volume, spatial refinement re-calibrates the output to highlight important locations within each such activated map. The *CDNetFW* employs the spatial and channel refinement mechanisms in a sequential manner, instead of applying them independently on the encoder volume to eventually combine the results as in [25]. Such sequential processing helps minimize redundant processing of irrelevant activation maps, while searching for spatial locations of interest within them. As a result, performance improvement occurs along with optimization of computational resources.

#### 3.3.1 Channel refinement

Consider the lower branch of the *FW* block at the top of Fig. 2. Let 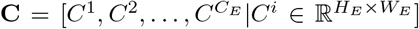 be the input volume from the encoder arm, where *C*_*E*_ indicates the number of encoder channels. Let *C*_*S*_ correspond to the number of decoder channels, 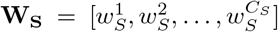 be the dilated convolution kernels, and 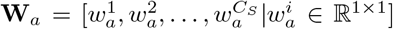 refer to the point-wise convolution kernels. The set of semantically rich activation maps 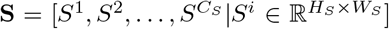 is passed through the *Assembled Dilated Convolution (AsDiC)* block, involving an assembly of convolutions with varying dilation rates *r* = 2, 3, 5 (as in Fig. 3). The *i*th dilated convolution 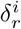 for kernel 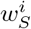 and feature map *f* ^*j*^, is expressed as

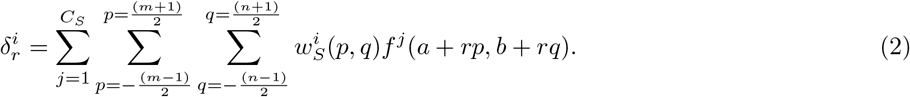

The application of multiple dilated convolutions on **S**, in a pipeline of the *AsDiC* block, yields the output feature map volume ***ν*** with component 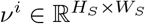 indicating the *i*th activation map. Hence

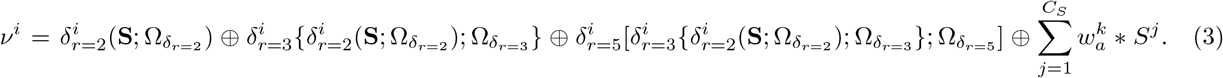

Here 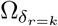 represents the set of parameters (at dilation rate *k*) defining the dilated convolutions **W**_**S**_, with *⊕* indicating element-wise addition, 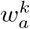 the *k*th point-wise convolution filter and *S*^*j*^ being the *j*th activation map in **S**. Use of multiple dilated convolutions, with varying dilation rates, produces multi-scalar views from the semantically rich activation map volume at deeper levels of *CDNetFW*. This helps capture the appropriate characteristics of the target COVID-19 lesions, which inherently exhibit inconsistent shapes and sizes. Use of point-wise convolution, along with multiple stacked dilated convolutions, help retain information from the regions overlooked by dilated convolutions. Larger field of view, without greater convolution kernel parameters, reduces the computational cost. Let us now focus on the lower branch of the *FW* block in Fig. 2. Depth-wise convolutions are applied to the output volume 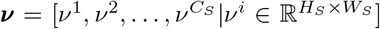 of *AsDiC* to squeeze its dimensions to 1 *×* 1 *× C*_*S*_, followed by a point-wise convolution for further dimensionality reduction in order to match the number of channels *C*_*E*_ of the encoder feature maps **C**. It introduces non-linearity to help learn more complex patterns. This is formulated as

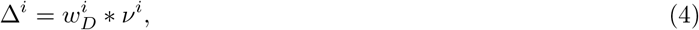

where 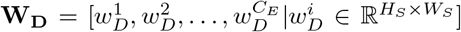 represent the depth-wise convolution kernels, and Δ^*i*^ *∈* ℝ ^1*×*1^ is the resultant *i*th output map. The derived weights *T* ^*i*^ *∈* [0, 1] are computed as

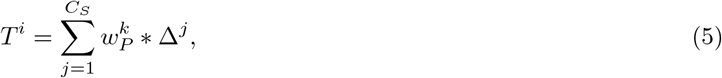

where 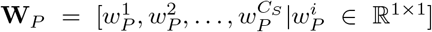 are the point-wise convolution kernel filters. The output is expressed as 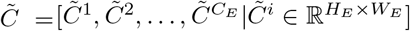

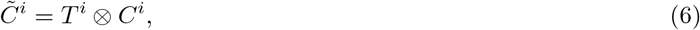

where *⊗* refers to the element-wise multiplication involving *C*^*i*^ (individual feature maps) of the encoder volume **C**. Unlike [25], which employed Global Average Pooling along with an artificial neural network to generate weights, this mechanism uses depth-wise convolutions. It aids independent learning of spatial patterns from each of the activation maps. This is important because each map is a representative of a distinct set of patterns. Besides, with such a fully convolutional module there exists no dependency on the dimensions of the input tensor; thus making the module flexible for incorporation in other architectures.

**Figure 2:**
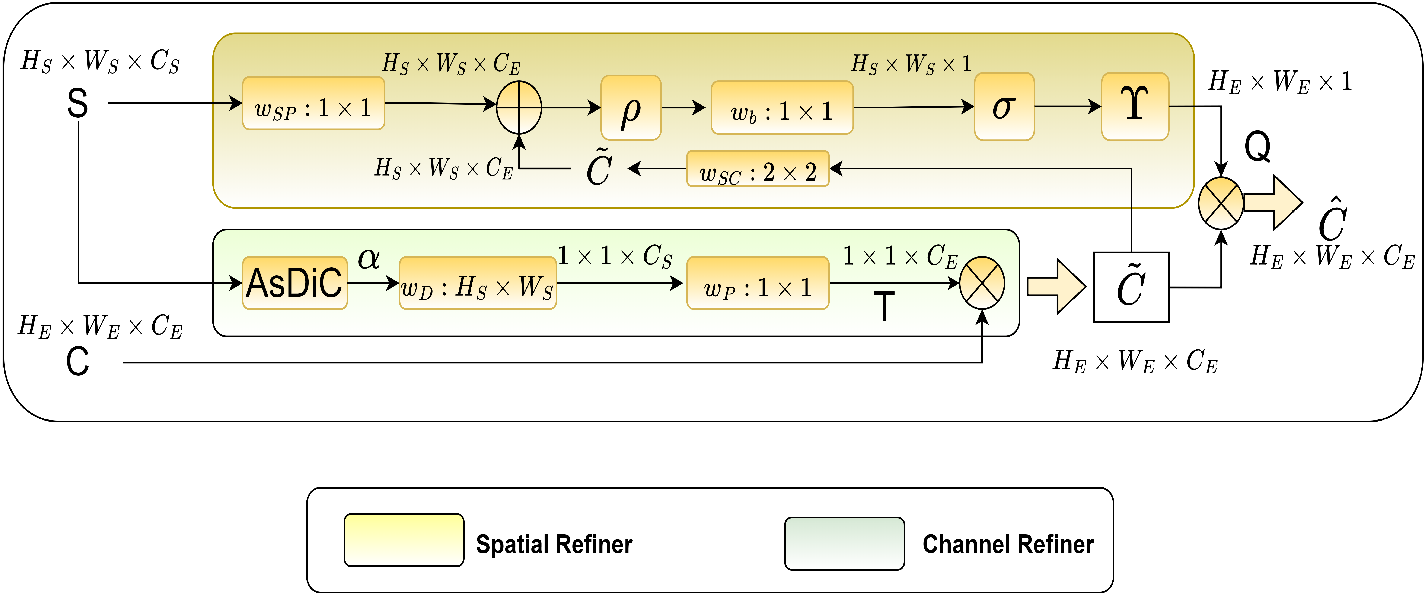
Feature Weighting module

**Figure 3:**
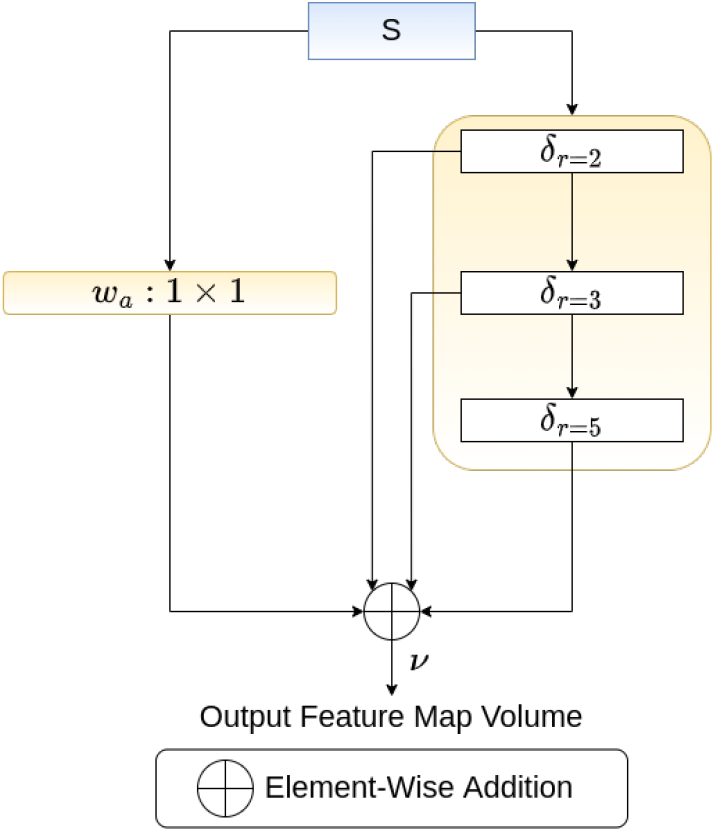
Assembled Dilated Convolution (*AsDiC*) kernels, involving different dilation rates

#### 3.3.2 Spatial refinement

The *i*th spatial-refinement weight *Q*^*i*^, illustrated in the upper branch of *FW* block in Fig. 2, is computed as

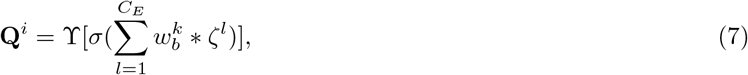

where

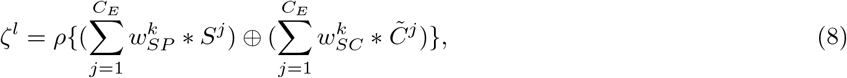

with 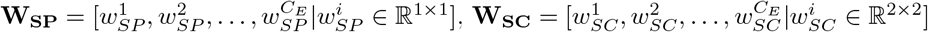 and 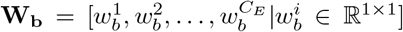 being the different convolution filters characterizing the spatial refinement mechanism. Here *ρ*(*x*) : max(0, *x*) and 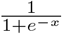 denote the ReLU and sigmoid activation functions, respectively, and ϒ represents the up-sampling operation using bilinear interpolation.

The spatially re-calibrated output features 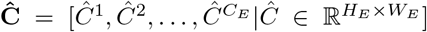 are obtained through an element-wise multiplication *⊗* of 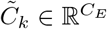, with the weight *Q*^*i*^ *∈* [0, 1] derived from the decoder signal volume **S**. We obtain

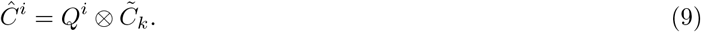

The formulation re-calibrates 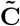 by the weights generated through the spatial refinement mechanism to produce signal **Ĉ**. This helps detect the relevant activation maps, from the entire volume, followed by focusing within them on only those locations carrying important information about the region of interest. It is unlike highlighting the relevant locations in the input encoder signal volume **C** with the generated weights 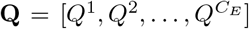 [25]. There is also reduction in the computational burden.

### 3.4 Loss functions

A significant problem encountered in medical image segmentation arises due to the severe class imbalance, often existing in a target region-of-interest (to be delineated) *vis-a-vis* the background region. This causes the final predictions of the model to be influenced by the dominant non-target (or background) regions. Dice loss [30] (*DL*) addresses the class imbalance issue between the foreground and background pixels. It is defined as

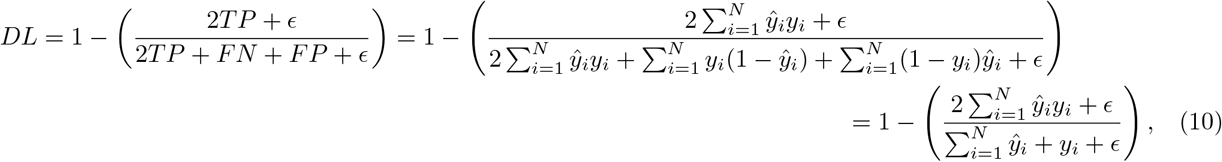

where *ŷ*_*i*_ and *y*_*i*_ are the predicted and ground truth value for the *i*th pixel, respectively, *N* is the total number of pixels, *ϵ* is a small random constant, *TP, FN, FP* correspond to the true positive, false negative, false positive, respectively. However, a significant disadvantage of this loss function is that it does not account for the output imbalance, encompassing *FP* and *FN*, in the segmentation output [31]. *Precision* specifies the proportion of pixels correctly identified as belonging to a given class (here, lesion region), relative to the total number of pixels annotated as representing that actual class. The percentage of correctly identified positive pixels (here, lesion) out of all positive pixels present in the ground truth, is represented by the metric *Recall*.

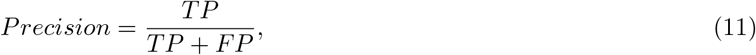

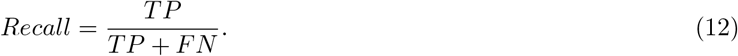

It is observed that training only with *DL* results in an increased *Precision* at the expense of diminished *Recall*. This is evident from the difference in the scores of *Precision* and *Recall* in Table 3.

The Tversky loss (*TL*) [32] alleviates the problem of *FP* and *FN* by assigning them with weight values *α* and *β*, respectively.

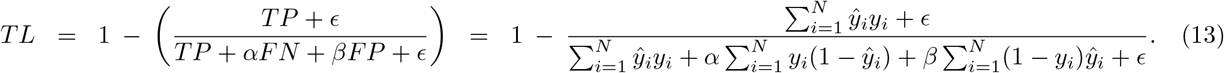

An enhanced version of *TL*, called Focal Tversky loss *FTL* [31], involves an additional parameter *γ* which helps target the imbalanced (smaller) Regions of Interest (ROIs) by exponentially increasing the loss for pixels with low predicted probability. This is expressed as

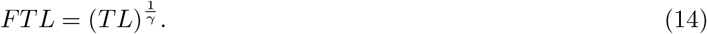

We used a linear combination *ℒ* of the loss functions *DL* and *FTL*, for training the proposed *CDNetFW*; thereby, utilizing the benefits of handling class imbalance while improving upon the balance between *Precision* and *Recall*. Here

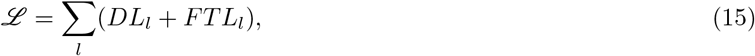

for *l* classes. Note that *l ∈* 0, 1 for binary segmentation. The total loss is computed as

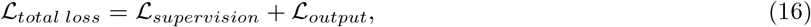

where

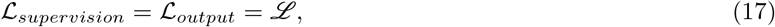

The values of the hyperparameters *α, β, γ*, were set at 0.7, 0.3, 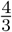, respectively [31].

### 3.5 Performance metrics

The *DSC, Precision, Recall, SPE*, and *IoU* are employed to evaluate the segmentation produced by the *CDNetFW*. The *DSC* is a harmonic mean of *Precision* and *Recall*, and measures the similarity between the predicted mask and the ground truth for a sample CT slice. The *SPE* measures the ability of the model to correctly identify pixels representing the background region (here, healthy lung tissues) out of all the pixels annotated as belonging to the background. Another important metric for the segmentation task is *IoU*, which calculates the amount of overlap between the predicted mask and the available ground truth. All these metrics have values lying in the range [0,1]. They are expressed as

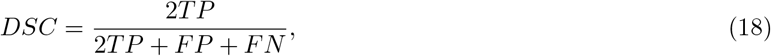

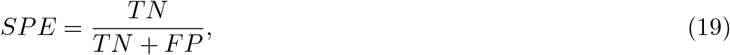

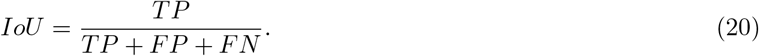

Here *TN* denotes *True Negative*. A classifier is represented in terms of a graphical plot between the *True Positive Rate (TPR)* and *False Positive Rate (FPR)*, at varying thresholds of classification. *Receiver Operating Characteristic (ROC)* curve is used as a comparative measure to evaluate between the classifiers. The Area Under the Curve (*AUC*) of each model is also a good evaluation measure. The model performance, under various loss functions, is depicted in terms of the Area Under the *Precision-Recall* curve *(AUC-PR)*.

## 4. Experimental Results

Here we outline the data sets used in the study, the implementation details, qualitative and quantitative experimental results, along with their comparative analysis.

### 4.1 Datasets

The COVID-19 lung CT slices were obtained from four publicly available datasets, as tabulated in Table 2. Dataset-1 [14] is a set of lung CT scans taken from 1110 patients belonging to five categories *viz*. CT-0, CT-1, CT-2, CT-3, CT-4. While CT-0 corresponds to the normal cases, the rest of the data is split into four groups according to increased severity of infection in the lung. Here CT-1 has 684 samples with affected lung percentage of 25% or below, CT-2 has 125 samples with affected area ranging from 25% to 50%, CT-3 consisting of 45 samples having 50% to 75% of affected lung region, and CT-4 encompassing just two samples with 75% and above affected lung portion. Only 50 scans, belonging to CT-1, were annotated with binary masks depicting regions of GGO. The affected lung area was assigned a label “1”, and the rest of the slice (unaffected portion without the lesion, as well as the background region) were assigned the label “0”. The CT volumes with annotated masks were used for our study. Dataset-2 [15] consists of 100 axial CT slices from *>* 40 COVID-positive patients. The slices were labelled by a radiologist to demarcate three different pathalogies of COVID-19 *viz*. GGOs, white consolidations and pleural effusion. Dataset-3 [16] encompasses nine volumetric lung CT scans obtained from the Italian Society of Medical Interventional Radiology. However, out of a total of 829 slices, only 373 slices are provided with annotations indicating the regions with GGOs and white consolidations. Dataset-4 [17] is a collection of lung CT scans from 20 patients, with annotations done by two radiologists which were later validated by another experienced radiologist. The ground truth of these slices consists of only two labels: “1” and “0” to indicate the diseased tissues and other regions (comprising healthy regions of lung and background). Here we used lung CT volumes from the first ten patients for extracting the slices in our experiments. This was because the remaining 10 samples contained non-uniform number of slices, being indicative of dissimilarity in the voxel spacing.

**Table 2:**
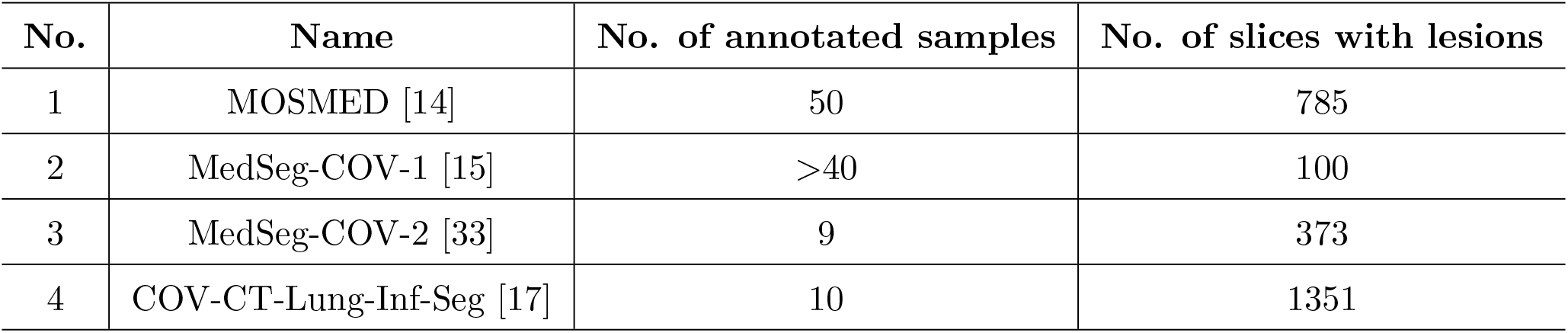
Datasets used

**Table 3:** Effect of loss functions on the performance metrics

| Loss functions | DSC | IoU | Precision | Recall |
| --- | --- | --- | --- | --- |
| Focal Tversky Loss | 0.8146 | 0.6964 | 0.7994 | **0.8386** |
| Dice Loss | 0.8217 | 0.7042 | 0.8593 | 0.7945 |
| **Focal Tversky Loss + Dice Loss** | **0.8293** | **0.7159** | 0.8372 | 0.8241 |
| Focal Loss + Dice Loss | 0.8192 | 0.70 | **0.8706** | 0.7799 |

All the slices were resized to a dimension of 512 *×* 512. The voxel intensities of all CT volumes, from the four data sources, were clipped to make them lie in the range [1000 *HU*, 170 *HU*], in order to filter out unnecessary details and noise. This was followed by intensity normalisation across the resultant multi-source database. Since all the CT slices in the volume do not contain COVID lesions, we selected only those having embedded lesions for the training. To account for the fact that not all datasets included labels for every possible COVID-19 pathology, we combined these different pathologies from Datasets 2 and 3 to create a single class representing COVID-19 lesions. The annotated samples, from the four multi-source datasets, were combined into a single database. This combined dataset was next randomly divided into five parts, for five-fold cross validation. Pooling the multi-source samples into one single dataset helped the model attain better generalization to learn the varying COVID-19 lesion structures and appearance (corresponding to different severity levels). We also utilized the additional ten patient lung masks from Dataset-4 to train the model for segmenting the lung region in the input CT slice while evaluating the severity of infection.

### 4.2 Results

The experimental setup for *CDNetFW* was implemented in Python 3.9 on a 12GB NVIDIA GeForce RTX 2080 Ti GPU. Optimizer Adam was used with an initial learning rate of 0.0001 for the first 35 epochs; which was subsequently reduced to 0.00001 for the remaining 65 epochs. Learning rate was reduced by a factor of 50%, when there was no further improvement in loss value after five consecutive epochs. Early stopping was employed to prevent overfitting. Both qualitative and quantitative analysis was made to evaluate the segmentation performance of the proposed *CDNetFW*. Comparative study with related state-of-the-art deep segmentation architectures demonstrate the superiority of our model, under different ablation experiments. The severity of infection was also estimated.

#### 4.2.1 Ablations

The effect of loss functions, *viz. FTL, DL, FL*+*DL, FTL*+*DL*, was investigated on the performance metrics of eqns. (11), (12, (18), (20), using the combined annotated dataset (as elaborated above). This is presented in Table 3. It is observed that *DSC* and *IoU* were the highest, when the proposed combined loss function of eqn. (15) was used [Row 3 in the table]. The number of pixels corresponding to infected lung region(s) is found to be significantly lower than the number of pixels from the background region, thereby resulting in major class imbalance. Fig. 4 corroborates that *AUC −PR* is the highest for the proposed combination, signifying the correct classification of a majority of the pixels. Even in scenarios involving smaller ROIs, it can achieve a better balance between *Precision* and *Recall*.

**Figure 4:**
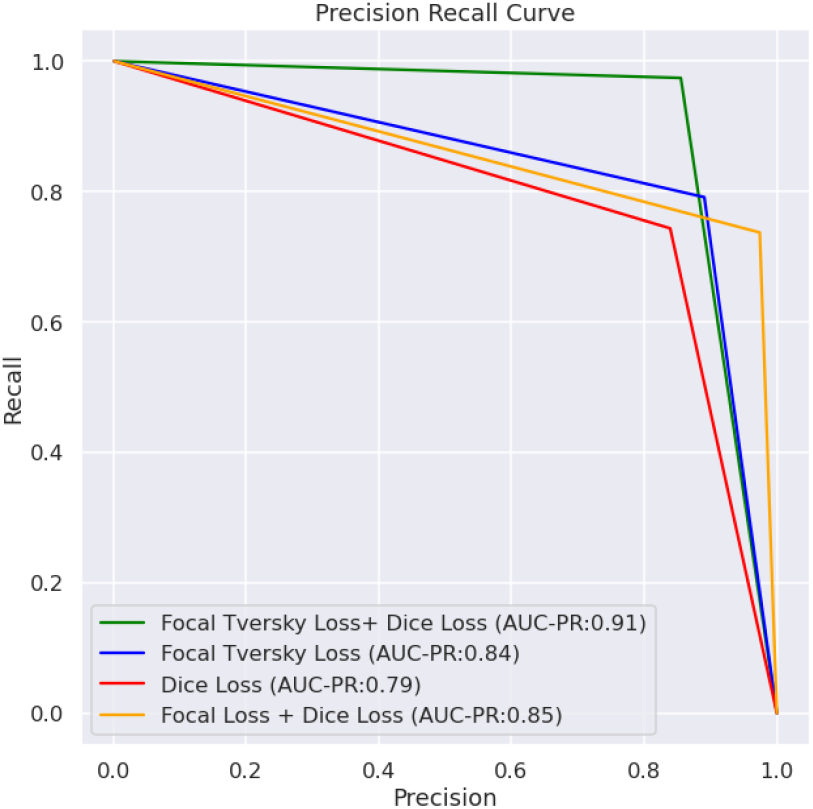
*Precision-Recall* curves for the different loss functions using the combined annotated dataset

Next some experiments were conducted to validate the role of *FW* and *AsDiC* blocks on the performance of the vanilla *U-*Net using the combined annotated dataset. It is observed from Table 4 that addition of *FW* block led to a rise in *DSC*. Upgrading the *FW* block with the *AsDiC* module resulted in a significant increase in the *Recall* score; implying a decrease in *FN* pixels. The role of feature weighting is further highlighted in Fig. 5 in terms of sample feature maps at the input and output of the *FW* block. It is evident that regions corresponding to the infected lung tissues get prominently displayed within the output feature maps (highlighted by yellow circles) in column (d) of the figure after passing through the *FW* module.

**Table 4:** Effect of different modules in *CDNetFW* using the combined annotated dataset

| Approach | $DSC$ | $Precision$ | $Recall$ | $SPE$ | $IoU$ |
| --- | --- | --- | --- | --- | --- |
| *U-Net* backbone + auxiliary branches + *FW* with *AsDiC* | **0.8145±0.014** | **0.8143±0.013** | **0.8216±0.0205** | **0.9976±0.0007** | **0.6962±0.0201** |
| *U-Net* backbone + auxiliary branches + *FW* w/o *AsDiC* | 0.8006±0.019 | 0.7984±0.025 | 0.8193±0.018 | 0.9975±0.0003 | 0.6787±0.026 |
| *U-Net* backbone + *FW* with *AsDiC* | 0.7698±0.033 | 0.7711±0.032 | 0.7909±0.0303 | 0.9971±0.0004 | 0.6408±0.0419 |
| *U-Net* backbone + *FW* w/o *AsDiC* | 0.7356±0.023 | 0.7631±0.084 | 0.7553±0.066 | 0.997±0.0009 | 0.5983±0.03 |
| *U-Net* backbone + auxiliary branches only | 0.7839±0.009 | 0.7941±0.022 | 0.7982±0.029 | 0.9976±0.0004 | 0.6594±0.0107 |
| Vanilla *U-Net* | 0.6306 ± 0.067 | 0.7821 ± 0.072 | 0.5916 ± 0.068 | 0.9969 ± 0.001 | 0.4873 ± 0.712 |

**Figure 5:**
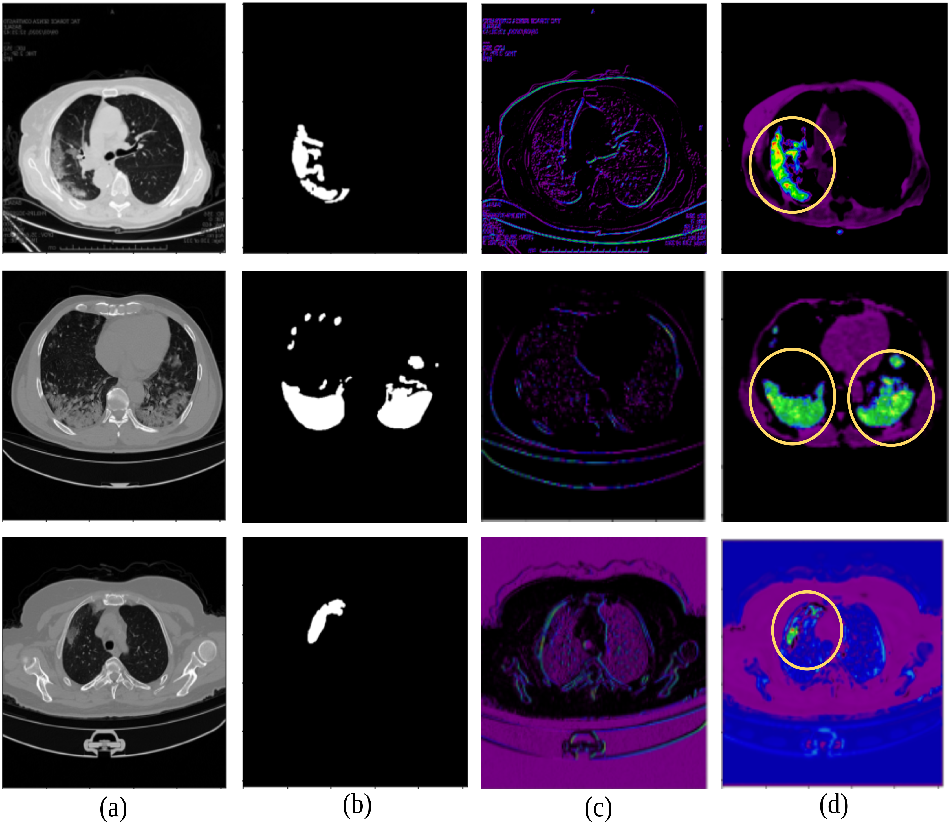
Impact of *FW* block, visualized with reference to (a) sample input CT slices, (b) corresponding ground truth, and the feature maps of *FW* block as displayed at its (c) input, & (d) output.

The incorporation of *auxiliary branches* to the basic *U-*Net framework resulted in an increment by a margin of *>*10 % in the *DSC* and *IoU*. A significant rise in both *Precision* and *Recall* are indicative of a simultaneous decrease in *FN* . Inclusion of *auxiliary branches*, in conjunction with the *FW* block, further enhanced the performance of *CDNetFW*; with precise segmentation, as expressed by highest *DSC* and *Recall* scores in Row 1 of Table 4.

Fig. 6 presents a visualization of the decrease in pixel count of *FN* pixels with respect to the segmentation output generated by the vanilla *U*-Net, as the different modules are added to the *CDNetFW*. It is observed that by incorporating both spatial and channel refinement mechanisms (in terms of *FW* and *AsDiC* modules), a better segmentation is obtained (both qualitatively and quantitatively) with reference to the ground truth. Adding auxiliary decoder branches generated the best outcome, as evident from part (f) of the figure.

**Figure 6:**
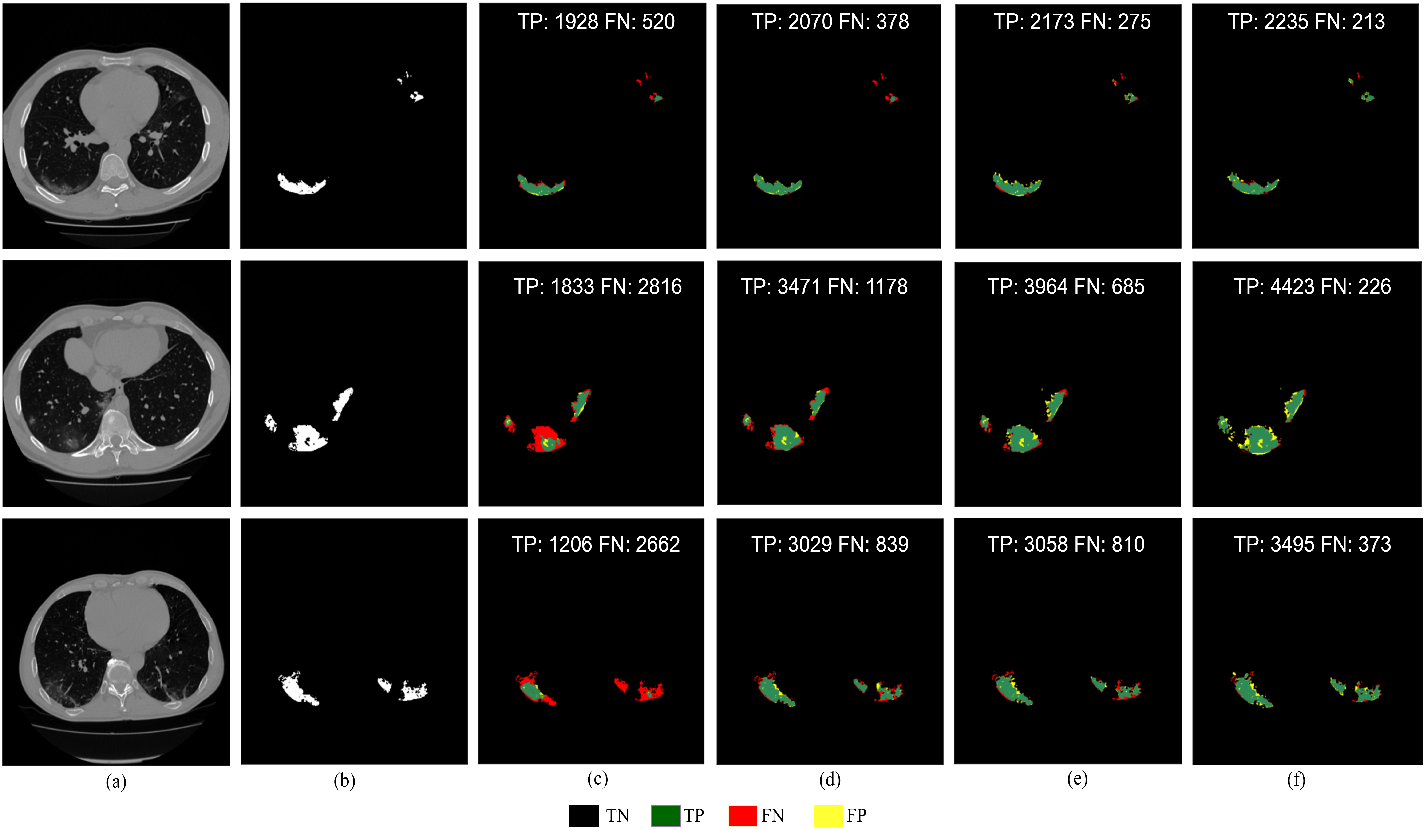
Segmentation on sample patients, using combined annotated dataset, depicting (a) input CT slice having (b) ground truth mask of lesions, and corresponding output generated by (c) vanilla *U*-Net, with incorporation of frequency refinement by (d) *FW* module *w/o AsDiC*, (e) *FW* module *with AsDiC*, and (f) auxiliary decoder branches.

**Figure 7:**
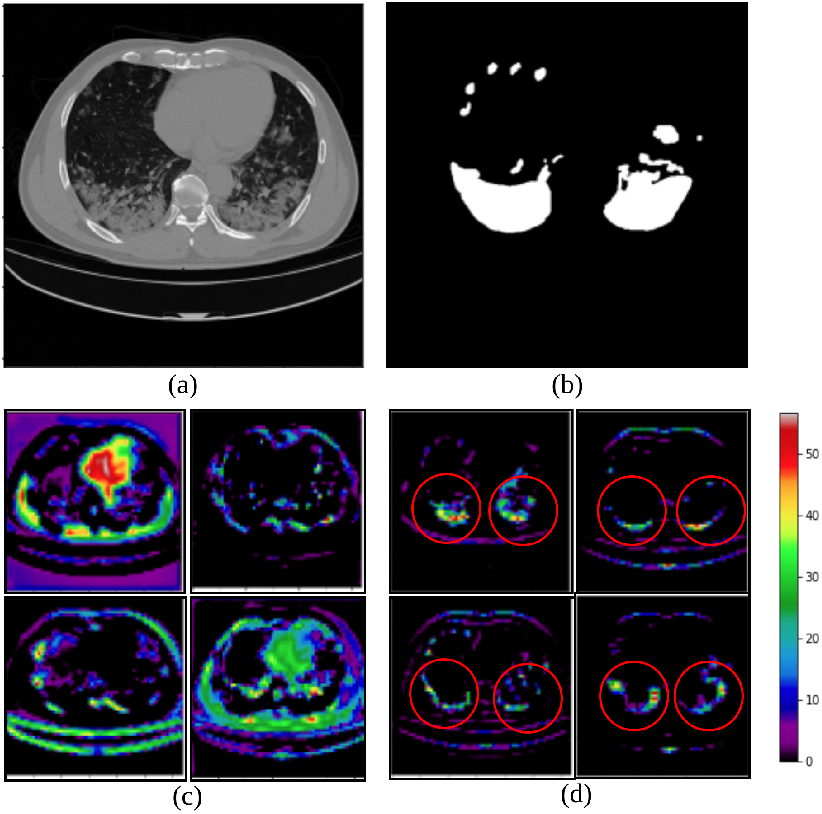
Feature maps from the fourth level of the encoder branch, using the combined annotated dataset. Sample (a) input CT slice, with (b) ground truth, and corresponding feature maps set (c) *without* auxiliary decoder branches, and (d) with auxiliary decoder branches

Inclusion of auxiliary decoder branches (at the encoder arm) generated feature map volumes containing activation maps, which focused on relevant patterns and features in the ROI, as observed from Fig. 7. The marked areas within the circles of part (d) of the figure are found to represent the extracted features corresponding to the anatomy of the lesions (with reference to the ground truth).

#### 4.2.2 Comparisons

Table 5 presents a comparative study of *CDNetFW* with state-of-the-art models, like *U-Net, U-Net++, Attention U-Net* and *Residual U-Net*, employing five-fold cross validation over the various performance metrics *DSC, Recall, IoU, Precision* and *SPE* (based on the combined annotated dataset). The best scores are marked in bold in the table. It is evident that *CDNetFW* outperforms the remaining models.

**Table 5:** Comparative study, over performance metrics, with related state-of-the-art architecture

| Models | *DSC* | *Precision* | *Recall* | *SPE* | *IoU* |
| --- | --- | --- | --- | --- | --- |
| *CDNetFW* | **0.8145±0.014** | **0.8143±0.013** | **0.8216±0.0205** | **0.9976±0.0007** | **0.6962±0.0201** |
| **Attention U-Net** [9] | 0.6587 ± 0.043 | 0.7537 ± 0.0751 | 0.6445 ± 0.038 | 0.9946 ± 0.006 | 0.52156 ± 0.041 |
| *U-Net* [8] | 0.6306 ± 0.067 | 0.7821 ± 0.072 | 0.5916 ± 0.068 | 0.9969 ± 0.001 | 0.4873 ± 0.712 |
| *U-Net++* [10] | 0.6881 ± 0.039 | 0.7209 ± 0.038 | 0.7083 ± 0.048 | 0.9961 ± 0.001 | 0.5456 ± 0.039 |
| **Residual U-Net** [11] | 0.6501 ± 0.018 | 0.7837 ± 0.047 | 0.6004 ± 0.044 | 0.9980 ± 0.0006 | 0.5123 ± 0.017 |

This reiterates the contention that by extending the vanilla *U-Net* framework with the novel feature weighting modules encompassing *FW* and *AsDiC*, along with the *auxiliary decoder branches*, enables *CDNetFW* to accurately identify the pixels belonging to the COVID-19 lesion category with a smaller number of *FN* s. By focusing only on the significant features that correspond to the target lesion regions, our architecture is found to be more effective in separating the target region(s) from a CT slice.

The values of *DSC* and *IoU*, the most widely used metrics of evaluation in medical image segmentation, demonstrate discernible improvement; thereby, establishing the efficacy of *CDNetFW*. Significant improvement in both *Precision* and *Recall* metrics indicate greater detection of *TP* pixels, with simultaneous reduction in the *FP* s. This validates the effectiveness of *CDNetFW* in capturing the structure of the infection lesions encompassing blurred edges with irregular contours and shapes.

Results are corroborated in terms of corresponding confusion matrices in Fig. 8. The box plot of *DSC* and the *ROC* curve with *AUC* in Fig. 9 additionally emphasize the superiority of our *CDNetFW*.

**Figure 8:**
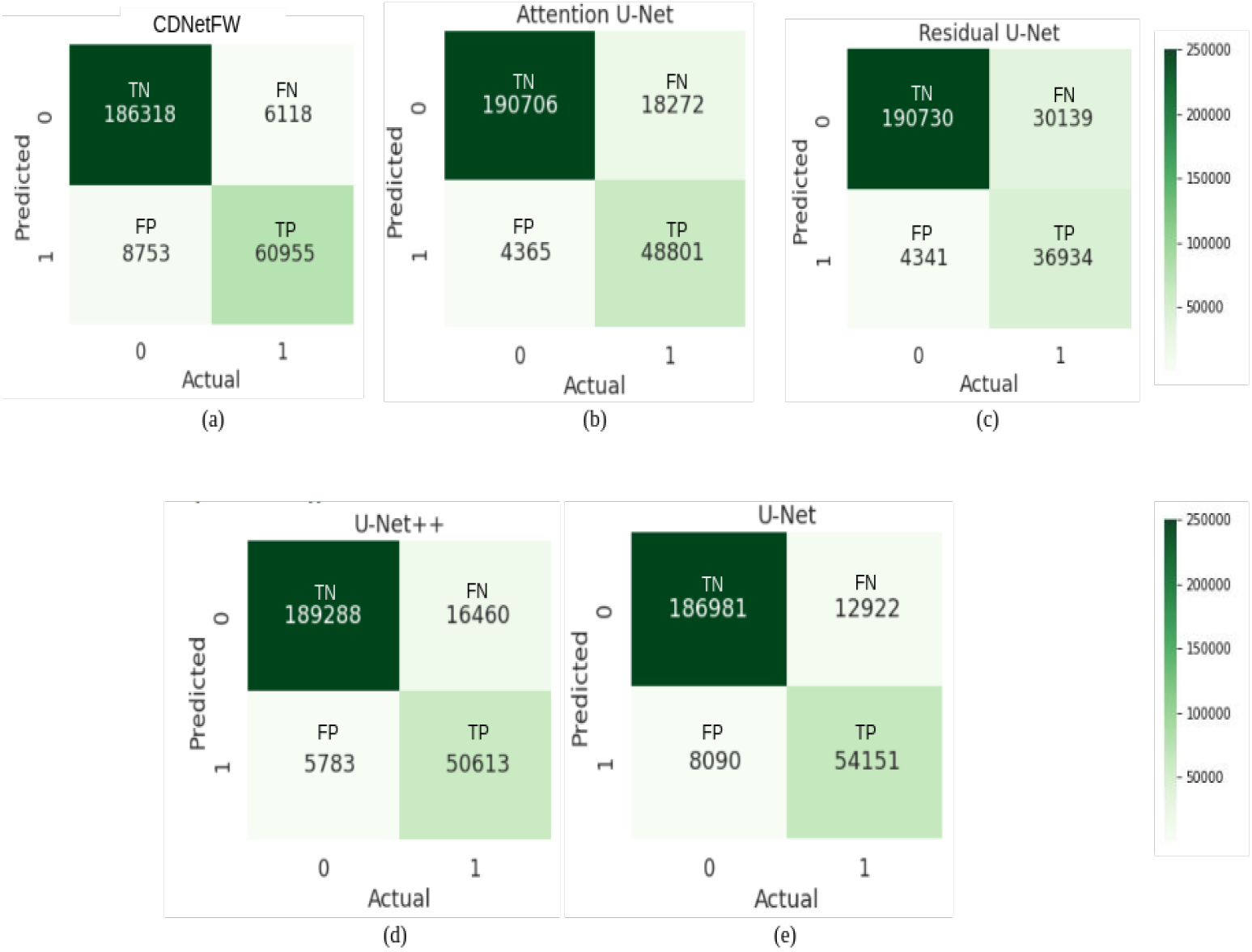
Confusion matrix, with combined annotated dataset, generated by (a) *CDNetFW*, (b) Attention *U*-Net, (c) Residual *U*-Net, (d)*U*-Net++, and (e) vanilla *U*-Net.

**Figure 9:**
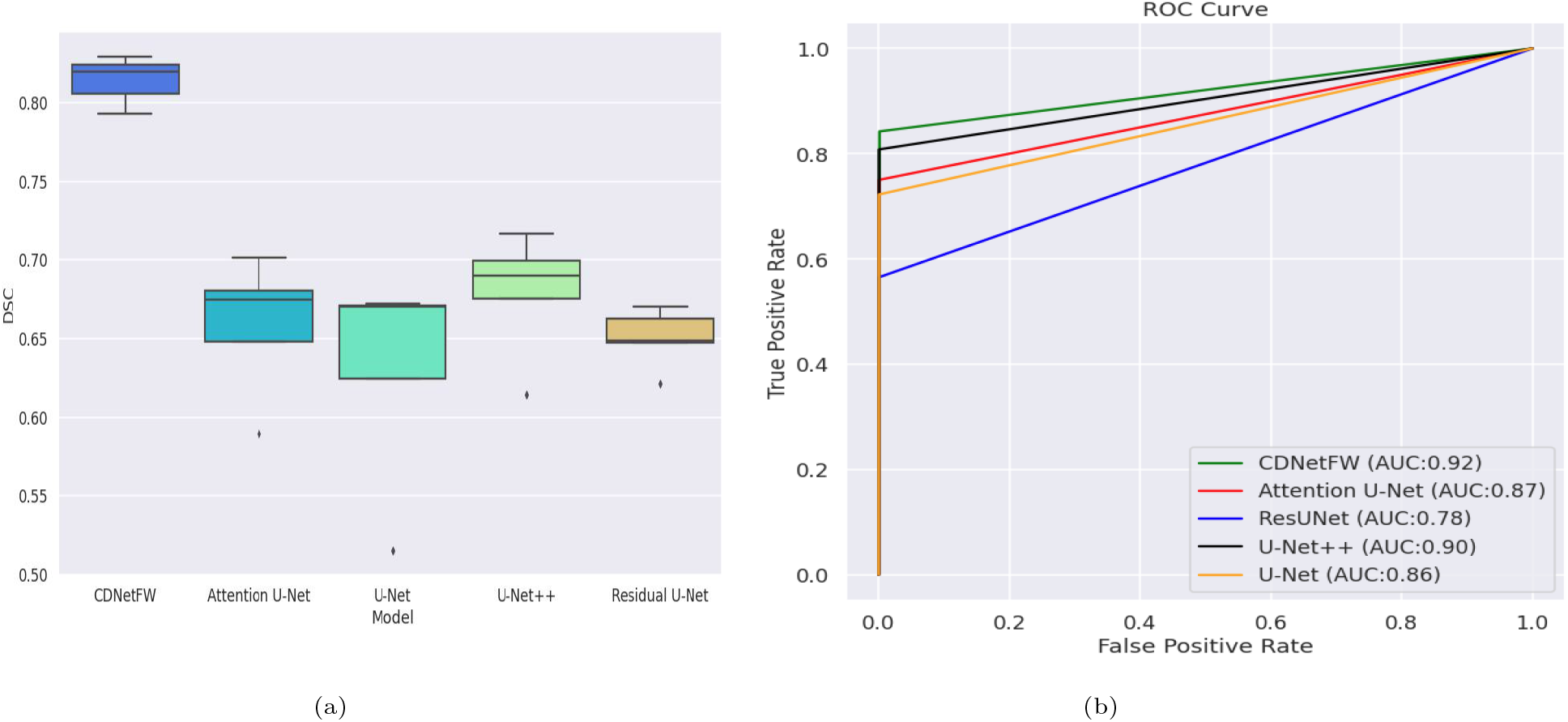
Comparative study with related models, on the combined annotated dataset, using (a) Box plot in terms of *DSC*, and (b) *ROC* curves with *AUC*.

Fig. 10 displays a qualitative study of the segmentation maps (on six sample CT scans), generated by our *CDNetFW*, and compared with respect to the related state-of-the-art models (referred above). It is evident that the vanilla *U*-Net, Attention *U*-Net, Residual *U*-Net, and *U*-Net++ are unable to extract the entire lesion region appropriately. Presence of *FN* pixels can be clearly viewed in the corresponding maps. On the other hand, the segmentation map of the *CDNetFW* indicates the highest concentration of *TP* pixels. It is visually evident that the *CDNetFW* outperforms the compared baseline architectures by more accurately segmenting the affected lung tissue, as observed from the sample CT slices.

**Figure 10:**
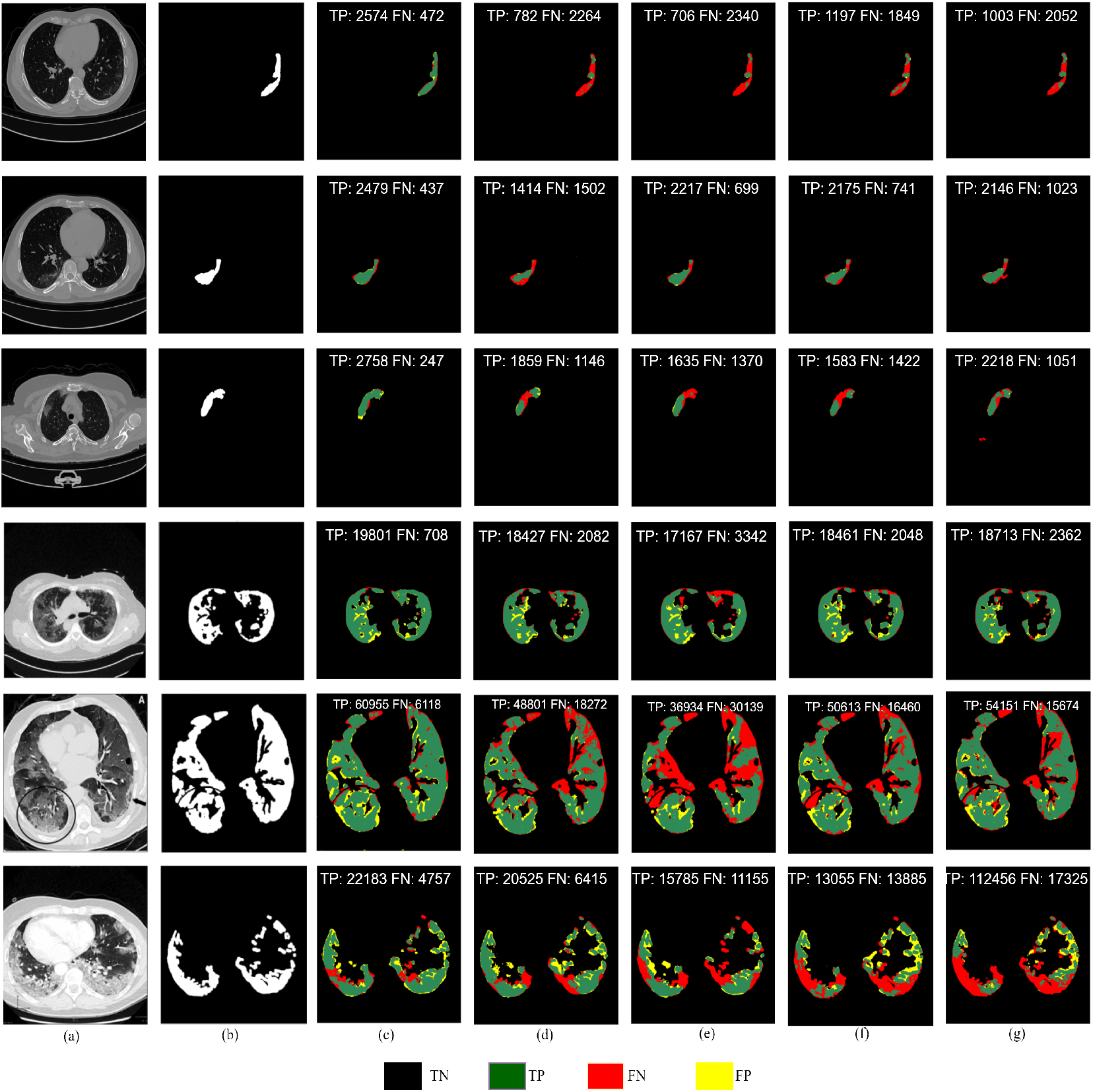
Qualitative comparison in segmentation output of *CDNetFW* with related state-of-the-art deep learning architectures, using the combined annotated dataset. (a) Sample CT slice, (b) the corresponding ground truth of the lesion, with predicted segmentation by (c) *CDNetFW*, (d) Attention *U −*Net (e) *U −*Net, (f) *U −*Net++, and (g) Residual *U −*Net.

Finally a comparative tabulation of a few other related state-of-the-art models (from literature), dealing with delineation of COVID lesions from lung CT, is provided in Table 6 in terms of *DSC*. The superiority of the proposed *CDNetFW* is observed.

**Table 6:** Comparative study with recent literature, in segmenting COVID lesions from lung CT

| Model | Characteristics | Dataset |  | DSC |
| --- | --- | --- | --- | --- |
|  |  | Train Set | Test Set |  |
| *CDNetFW* | Composite deep network, with feature weighting | 80% of CT images from MOSMED [14], MedSeg-COV-1 [15], MedSeg-COV-2 [33] and COV-CT-Lung-Inf-Seg [17] | 20% of CT images from MOSMED [14], MedSeg-COV-1 [15], MedSeg-COV-2 [33] and COV-CT-Lung-Inf-Seg [17] | **0.82** |
| *Inf-Net* | Parallel partial decoder to generate coarse output, with refinement by reverse and edge attention | 45 randomly selected CT images from MedSeg-COV-1 [15] | 50 CT images from MedSeg-COV-1 [15], | 0.682 |
| Goncharov et al | Multi-task approach for segmentation with classification | MOSMED [14], MedSeg-COV-2 [33], COV-CT-Lung-Inf-Seg [17] | MOSMED [14] | 0.63 |
| *D2A-Net* | Dual-attention and hybrid dilated convolutions | MedSeg-COV-2 [33], COV-CT-Lung-Inf-Seg [17] | MedSeg-COV-1 [15] | 0.72 |
| *nCoVSegNet* | Transfer learning, channel and spatial attention | 40 cases from MOSMED [14], LIDC-IDRI | 10 cases from MOSMED [14], COV-CT-Lung-Inf-Seg [17] | 0.69 |
| *LCOV-Net* | Lightweight CNN with attention | 80% of CT images from 10 private hospitals | 20% of CT images from 10 private hospitals | 0.78 |

#### 4.2.3 Severity of infection

The severity grading of a sample patient was computed, based on the infection region in the corresponding lung CT. The predicted lung mask and the lesion segmentation mask were used to calculate the ratio of the affected lung area to total lung area, individually for the left and right lung. The maximum of these two ratios determined the grade of severity of infection in the patient [2]. The severity grades are assigned according the following criteria: (i) CT-0: Healthy patients (ii) CT-1: Patients with infected lung % *<*25 (iii) CT-2: Patients with infected lung % between 25-50 (iv) CT-3: Patients with infected lung % between 50 and 75 (v) CT-4: Patients with infected lung % *>*75 Fig. 11 depicts the sample lung CT slices of five different patients, highlighting their maximum visible lung area with reference to the total CT volume in each case. The ratio computation results in the following prediction.

**Figure 11:**
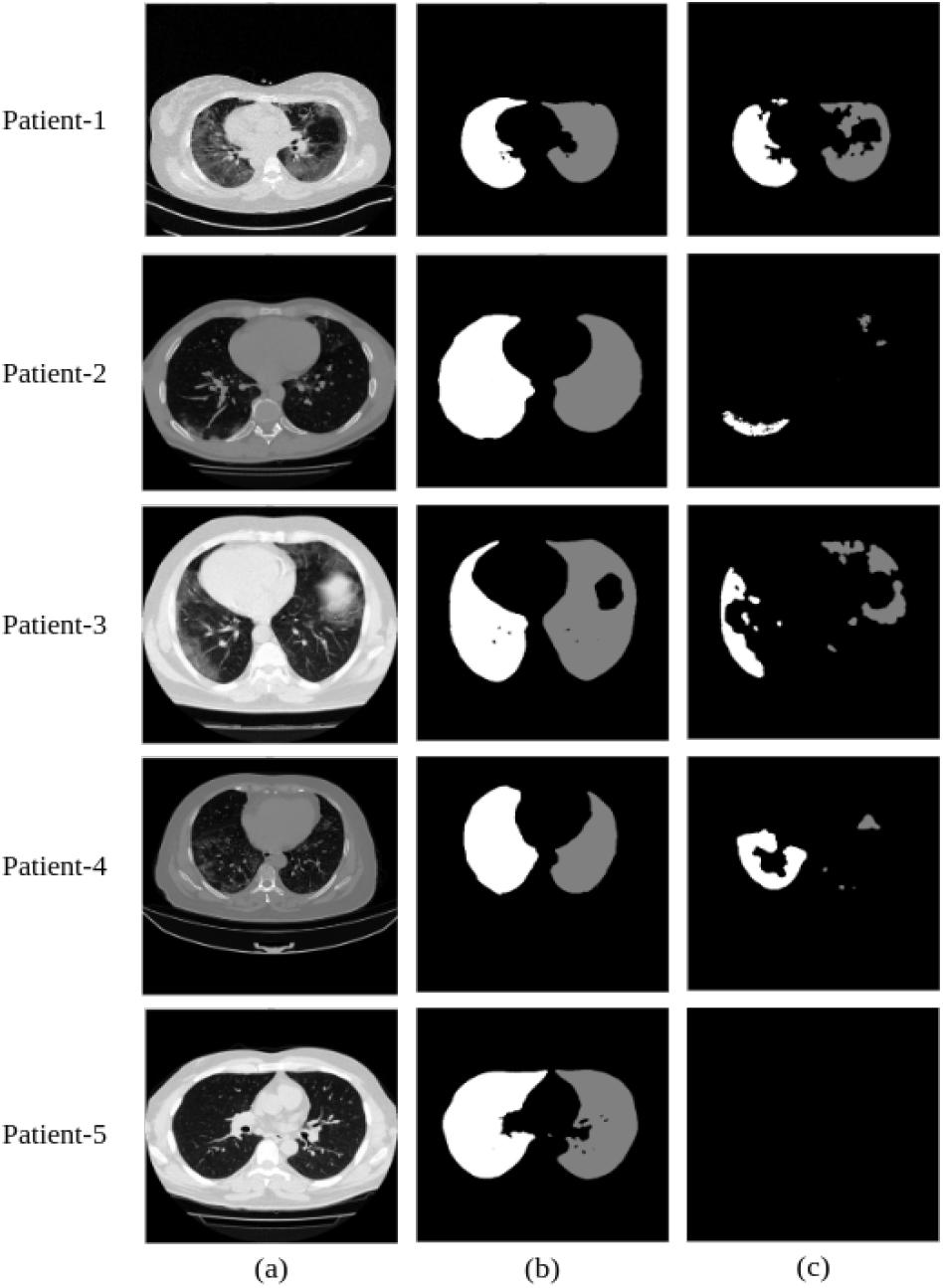
Gradation of severity in sample COVID-infected patients, depicting the (a) lung CT slice, (b) lung mask (white and gray indicating left and right lung, respectively), and the corresponding delineated lesion regions from both lungs.

- Patient-1: Affected Left Lung % = 81.92, Affected Right Lung % = 69.00, Grade: CT-4;
- Patient-2: Affected Left Lung % = 7.75, Affected Right Lung % = 2.27, Grade: CT-1;
- Patient-3: Affected Left Lung % = 31.00, Affected Right Lung % = 23.06, Grade: CT-2;
- Patient-4: Affected Left Lung % = 38.36, Affected Right Lung % = 6.40, Grade: CT-2;
- Patient-5: Affected Left Lung % = Nil, Affected Right Lung % = Nil, Grade: CT-0.

## 5. Conclusions and Discussion

A novel composite deep architecture *CDNetFW*, which integrates a mini-segmentation network with feature weighting mechanism, has been developed for the accurate segmentation of COVID lesions from lung CT slices. The *FW* and *AsDiC* modules contribute towards feature refinement for effective delineation of irregular shapes and contours of lesions, their low contrast, and associated blurring. The network allows highlighting important activation maps, while focusing on key locations within them; thereby, allowing *CDNetFW* to accurately delineate the relevant lesions. Auxiliary branches enable learning of stronger and more discriminating features at shallower levels of the network. This enhances the output segmentation quality. Class imbalance and false negatives are tackled using a combination loss, while outlining the lesion structure. This also leads to accurate computation of the affected lung area to help grade the severity of infection.

In the absence of sufficient annotated samples, the model could not be trained to distinguish between the different pathologies, like GGO, white consolidation, and pleural effusion. The research is currently being extended to multi-class segmentation. It can result in an effective tool for the screening and early detection of patients who may have contracted the COVID-19 pathogen. In individual patients infected with the virus, and demonstrating associated pulmonary abnormalities, the same methodologies can be used to accurately and more rapidly assess disease progression and guide therapy with effective patient management. Volumetric modeling of the lung can help in determining the optimal prognostics for recovery, based on the existing damage. Incorporation of additional genomic inputs is expected to throw further illumination on the investigation.

## Data Availability

All data sources in the present work are contained in the manuscript

## Acknowledgments

This work was supported by the J. C. Bose National Fellowship, sanction no. JCB/2020/000033 of S. Mitra.

## References

[1] O. Gozes, M. Frid-Adar, and et al., “Rapid AI development cycle for the Coronavirus (COVID-19) pandemic: Initial results for automated detection & patient monitoring using deep learning CT image analysis,” arXiv:2003.05037, 2020. [Online]. Available: https://spectrum.ieee.org/hospitals-deploy-ai-tools-detect-covid19-chest-scans

[2] S. P. Morozov, V. Y. Chernina, and et al., “Chest computed tomography for outcome prediction in laboratory-confirmed COVID-19: A retrospective analysis of 38,051 cases,” Digital Diagnostics, vol. 1, no. 1, pp. 27–36, 2020.

[3] Y. LeCun, Y. Bengio, and et al., “Deep learning,” Nature, vol. 521, pp. 436–444, 2015.

[4] K. He, X. Zhang, and et al., “Deep residual learning for image recognition,” in Proceedings of the IEEE Conference on Computer Vision and Pattern Recognition, 2016, pp. 770–778.

[5] S. H. Gao, M. M. Cheng, and et al., “Res2Net: A new multi-scale backbone architecture,” IEEE Transactions on Pattern Analysis and Machine Intelligence, vol. 43, pp. 652–662, 2019.

[6] G. Huang, Z. Liu, and et al., “Densely connected convolutional networks,” in Proceedings of the IEEE Conference on Computer Vision and Pattern Recognition, 2017, pp. 4700–4708.

[7] V. Badrinarayanan, A. Kendall, and et al., “SegNet: A deep convolutional encoder-decoder architecture for image segmentation,” IEEE Transactions on Pattern Analysis and Machine Intelligence, vol. 39, pp. 2481–2495, 2017.

[8] O. Ronneberger, P. Fischer, and et al., “U-Net: Convolutional networks for biomedical image segmentation,” in Proccedings of the International Conference on Medical Image Computing and Computer Assisted Intervention, (MICCAI). Springer, 2015, pp. 234–241.

[9] O. Oktay, J. Schlemper, and et al., “Attention U -Net: Learning where to look for the pancreas,” in Proceedings of the Medical Imaging with Deep Learning, 2018.

[10] Z. Zhou, R. Siddiquee, and et al., “UNet++: A nested U-Net architecture for medical image segmentation,” in Deep Learning in Medical Image Analysis and Multimodal Learning for Clinical Decision Support. Springer, 2018, pp. 3–11.

[11] A. Khanna, N. D. Londhe, and et al., “A deep Residual U-Net convolutional neural network for automated lung segmentation in computed tomography images,” Biocybernetics and Biomedical Engineering, vol. 40, no. 3, pp. 1314–1327, 2020.

[12] V. S. Tseng, J. J. C. Ying, and et al., “Computational intelligence techniques for combating COVID-19: A survey,” IEEE Computational Intelligence Magazine, vol. 15, pp. 10–22, 2020.

[13] J. S. Suri, S. Agarwal, and et al., “A narrative review on characterization of acute respiratory distress syndrome in COVID-19-infected lungs using artificial intelligence,” Computers in Biology and Medicine, vol. 130, p. 104210, 2021. [Online]. Available: https://doi.org/10.1016/j.compbiomed.2021.104210

[14] S. P. Morozov, A. E. Andreychenko, and et al., “MOSMED data: Data set of 1110 chest CT scans performed during the COVID-19 epidemic,” Digital Diagnostics, vol. 1, pp. 49–59, 2020.

[15] MedSeg, H. B. Jenssen, and et al., “MedSeg Covid Dataset 1,” 1 2021. [Online]. Available: https://figshare.com/articles/dataset/MedSegCovidDataset1/13521488

[16] MedSeg, H. B. Jenssen, and et al., “MedSeg COVID Dataset 2,” 2021. [Online]. Available: https://figshare.com/articles/dataset/CovidDataset2/13521509

[17] J. Ma, C. Ge, and et al., “COVID-19 CT Lung and Infection Segmentation Dataset,” Apr. 2020. [Online]. Available: https://doi.org/10.5281/zenodo.3757476

[18] L. Li, L. Qin, and et al., “Artificial intelligence distinguishes COVID-19 from community acquired pneumonia on chest CT,” Radiology, vol. 296, 2020. [Online]. Available: https://doi.org/10.1148/radiol.2020200905

[19] S. A. Harmon, T. H. Sanford, and et al., “Artificial intelligence for the detection of COVID-19 pneumonia on chest CT using multinational datasets,” Nature Communications, vol. 11, 2020. [Online]. Available: https://doi.org/10.1038/s41467-020-17971-2

[20] Y. H. Wu, S. H. Gao, and et al., “JCS: An explainable COVID-19 diagnosis system by joint classification and segmentation,” IEEE Transactions on Image Processing, vol. 30, pp. 3113–3126, 2021. [Online]. Available: https://doi.org/10.1109/TIP.2021.3058783

[21] Z. Han, B. Wei, and et al., “Accurate screening of COVID-19 using attention-based deep 3D multiple instance learning,” IEEE Transactions on Medical Imaging, vol. 39, pp. 2584–2594, 2020.

[22] A. Saood and I. Hatem, “COVID-19 lung CT image segmentation using deep learning methods: U -Net versus SegNet,” BMC Medical Imaging, vol. 21, pp. 1–10, 2021.

[23] M. Goncharov, M. Pisov, and et al., “CT-Based COVID-19 triage: Deep multitask learning improves joint identification and severity quantification,” Medical Image Analysis, vol. 71, p. 102054, 2021. [Online]. Available: https://doi.org/10.1016/j.media.2021.102054

[24] F. Xie, Z. Huang, and et al., “DUDA-Net: A double U-shaped dilated attention network for automatic infection area segmentation in COVID-19 lung CT images,” International Journal of Computer Assisted Radiology and Surgery, vol. 16, pp. 1425–1434, 2021.

[25] X. Zhao, P. Zhang, and et al., “D2A U -Net: Automatic segmentation of COVID-19 CT slices based on dual attention and hybrid dilated convolution,” Computers in Biology and Medicine, vol. 135, p. 104526, 2021. [Online]. Available: https://doi.org/10.1016/j.compbiomed.2021.104526

[26] J. Liu, B. Dong, and et al., “COVID-19 lung infection segmentation with a novel two-stage cross-domain transfer learning framework,” Medical Image Analysis, vol. 74, p. 102205, 2021.

[27] J. Deng, W. Dong, and et al., “ImageNet: A large-scale hierarchical image database,” in Proceedings of IEEE Conference on Computer Vision and Pattern Recognition, 2009, pp. 248–255.

[28] D. P. Fan, T. Zhou, and et al., “Inf-Net: Automatic COVID-19 lung infection segmentation from CT images,” IEEE Transactions on Medical Imaging, vol. 39, pp. 2626–2637, 2020.

[29] Q. Zhao, H. Wang, and et al., “LCOV-NET: A lightweight neural network for COVID-19 pneumonia lesion segmentation from 3D CT images,” in Proceedings of the IEEE 18th International Symposium on Biomedical Imaging (ISBI), 2021, pp. 42–45.

[30] F. Milletari, N. Navab, and et al., “V-net: Fully convolutional neural networks for volumetric medical image segmentation,” in Proceedings of Fourth International Conference on 3D vision (3DV). IEEE, 2016, pp. 565–571.

[31] N. Abraham and N. M. Khan, “A novel focal Tversky loss function with improved attention U-Net for lesion segmentation,” in Proceedings of IEEE 16th International Symposium on Biomedical Imaging (ISBI 2019), 2019, pp. 683–687.

[32] S. S. M. Salehi, D. Erdogmus, and et al., “Tversky loss function for image segmentation using 3D fully convolutional deep networks,” in Proceedings of International Workshop on Machine Learning in Medical Imaging. Springer, 2017, pp. 379–387.

[33] “COVID-19 CT Segmentation Dataset,” 2020. [Online]. Available: http://medicalsegmentation.com/covid19/

